# Sexual health at the end of life in patients with advanced cancer and their partners. Results of a Dutch prospective longitudinal study (eQuiPe)

**DOI:** 10.1101/2025.07.03.25330800

**Authors:** Isabel S. van der Meer, Moyke A.J. Versluis, Heidi P. Fransen, Corien Eeltink, Arnold Baars, Dirkje Sommeijer, Tineke Smilde, Annemieke van der Padt-Pruijsten, Lonneke V. van de Poll-Franse, Natasja J.H. Raijmakers

## Abstract

**Background:** Sexual health can be negatively affected by cancer. There is a paucity of literature exploring changes in sexual health during the end of life.

**Aim:** To assess changes in sexual health in patients with advanced cancer and their partners at the end of life, and to identify the associated factors.

**Design:** A prospective longitudinal study of patients with advanced cancer and their partners on the quality of care and quality of life (QoL) (eQuiPe). Patients and partners completed 3-monthly questionnaires. Sexual health (desire, activity, satisfaction and enjoyment) was measured using the EORTC QLQ-SH22.

**Setting/participants:** Patients aged ≥18 years and diagnosed with advanced cancer were recruited in one of the forty participating Dutch hospitals. Relatives were recruited through patients, and for this study only couples (patient-partner) were included (n=352).

**Results:** Towards death, patients remained relatively stable in sexual activity (range 15-19), satisfaction (range 40-45), and enjoyment (range 30-44). Similar results were found for partners. In patients, sexual desire significantly decreased towards the end of life (β 0.4, 95%CI 0.1-0.7). Moreover, greater decline in physical functioning was associated with poorer outcomes in most aspects of sexual health. Sexual desire, activity and satisfaction were individually associated with QoL in patients.

**Conclusions:** Sexual health remains relatively stable at the end of life in patients with advanced cancer and their partner. Patients with worse physical functioning report worse sexual health and sexual desire, activity and satisfaction are individually associated with better QoL. Therefore, addressing sexual health in palliative care is essential.

## Background

In the Netherlands approximately 38,000 people are diagnosed with advanced cancer each year (1). Despite significant medical advances in oncological care, patients with advanced cancer still have a limited median survival (2,3). The diagnosis and treatment of advanced cancer have a significant impact on patients and their relatives and maintaining or improving the quality of life (QoL) for these patients is paramount.

Sexual health is an integral part of QoL and consists of several components, including sexual activity, enjoyment, satisfaction, and intimacy (4). Advanced, incurable cancer and subsequent treatments can have negative implications for sexual health, both for patients as their partners. Over 40% of patients with curable cancer encounter sexual problems post-treatment (5,6). The majority of patients with advanced cancer (75%) reported low sexual satisfaction (7) and lower levels of sexual activity (8). Moreover, patients with advanced cancer reported lower sexual health compared to patients with curable cancer, due to side effects of treatment and physical changes at the end of life (9).

The dynamics of sexual health and its relation to QoL of patients with advanced cancer may change throughout the disease trajectory, as most patients reported fluctuations in the frequency and intensity of sexuality, especially in the last months of life (7,9,10). Changes in sexual health emerge from physical, mental and emotional transformations, such as changes in body image, and shifts in relationship dynamics due to altered social roles (11,12). However, the importance of sexuality for patients with advanced cancer remains relatively unchanged at the end of life (10,13).

Current literature consists mainly of cross-sectional studies on sexual health, with a limited understanding of the changes in sexuality during at the end of life (8,10,13,14). Additionally, the majority of sexual health studies have focused on sexual functioning (6,9,15–18) and on cancer survivors (12,17,19–21), with an emphasis on female patients (16,21,22–26). Understanding how sexual health changes in the end of life is needed to support patients and their partners to optimize QoL. Such knowledgde is essential to raise awareness among healthcare providers of the importance of sexual health in the management of advanced cancer. Therefore, the aim of this study was to assess the changes in sexual health among adult patients with advanced cancer and their partners in the end of life, and the factors associated with these changes.

## Methods

### Study design and ethics

A longitudinal, multicenter, prospective, observational cohort study (eQuiPe) was conducted to assess the QoL and quality of care among patients with advanced cancer and their relatives in the Netherlands (27). Patients were approached either through direct engagement by the attending physician at one of the 40 hospitals, or through self-registration between November 2017 and March 2020. Initial contact with patients was established via telephone by the research team, with inquiries made regarding the willingness of relatives to join the study. Written informed consent was a prerequisite for study participation. Patients and relatives received a baseline questionnaire followed by subsequent three-monthly follow-up questionnaires until the patient’s death. Questionnaires were administered either online or on paper via the Patient Reported Outcomes Following Initial treatment and Long-term Evaluation of Survivorship (PROFILES)(28). Clinical data of patients were obtained by linking to the to the Netherlands Cancer Registry (NCR)(29).

The eQuiPe study was exempted from full medical ethical review by the Antoni van Leeuwenhoek’s hospital Medical Ethics Committee (METC17.1491), in accordance with the Dutch Medical Research Involving Human Subjects Act (WMO). Registration details of the study can be accessed through the Netherlands Trial Register under the identifier NTR6584, and additional information on the study has been published elsewhere (27).

### Study population

Patients and relatives had to be aged 18 years and older and able to complete the questionnaire in Dutch to be deemed eligible for inclusion in the study. Additional information can be found elsewehere (Appendix 1). For this analysis, deceased patients of whom a partner also completed a questionnaire in the last 18 months of the patient’s life were selected. In cases where neither the patients nor their partners responded to any of the sexual health questions, both the patient and the partner were excluded from the final analysis. Of the 566 couples, 352 couples were included in the final analysis (Figure 1).

**Figure 1.**
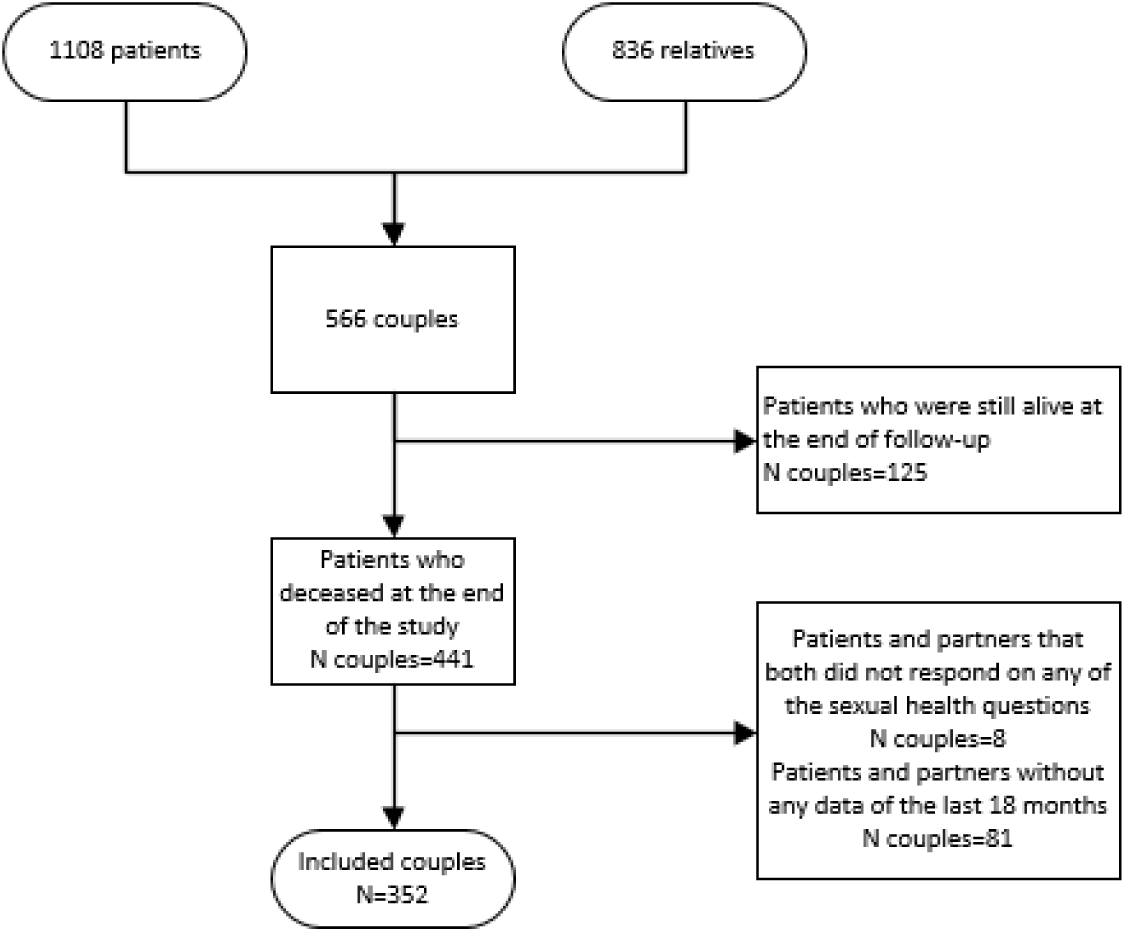
Flowchart of the study procedure.

### Measures

#### Sexual health

Sexual health was assessed at baseline and three-monthly follow-up questionnaire using the European Organization for Research and Treatment of Cancer Quality of Life Questionnaire Sexual Health (EORTC QLQ-SH22) (30,31). The sexual health subscale comprised four questions and related to the past four weeks, measuring sexual desire, sexual activity, sexual satisfaction, and sexual enjoyment. Response options included a 4-point Likert scale ranging from 1 ‘Not at all’ to 4 ‘Very much’. Responses were linearly transformed to a scale of 0-100, with higher scores indicating better outcomes (30,31).

#### Socio-demographic and clinical characteristics

Age, sex, religion, education level, relationship duration, having children, and children living at home were self-reported in the baseline questionnaire. Education was categorized according to the International Standard Classification of Educational Guidelines: low, middle and high (32). Patients and partners/relatives were asked to which religion they belonged, which was categorized in being religious or not. Comorbidities were measured at baseline using the Self-administered Comorbidity Questionnaire (SCQ) (31). Primary tumor type and date of death were obtained from the NCR (29). Time to death was defined as the time (in months) between completion of the questionnaire and date of death. Subsequently, in each questionnaire, patients reported if they received cancer treatment (including chemotherapy, radiotherapy, immunotherapy, hormonal therapy or other therapies) in the last 18 months of life and was eventually incorporated as a binary variable (yes/no).

#### Body image

Body image was measured by the validated Body Image Scale (BIS), with higher scores indicating a lower body image. The BIS comprised of ten questions and measured the perception of their body image in the past week, with responses standardized on a 0-30 scale (33).

#### Quality of life

Physical functioning and global QoL were measured by the European Organization for Research and Treatment of Cancer Quality of Life Questionnaire C30 (EORTC QLQ-C30) (34). For both physical functioning and global QoL, responses were linearly transformed to a 0-100 score, with higher scores indicating better functioning/higher global QoL.

## Statistical analysis

Descriptive statistics were used to summarize the socio-demographic and clinical characteristics of patients and their partners. Time to death was categorized into cohorts of 3 months: 16-18 months, 13-15 months, 10-12 months, 7-9 months, 4-6 months, and 0-3 months before death to summarize the course of sexual desire, activity, satisfaction, and enjoyment over time. Mixed-effects multivariable linear regression was performed to assess changes in sexual health over the last 18 months of the patient’s life for both patients and partners separately. The analysis among patients included time to death (continuous), age, sex, body image, treatment, comorbidities, physical functioning, treatment, tumor type, QoL, and QoL of the partner (11,35,36). The regression of the partner also included the patient’s time to death, treatment, primary tumor and QoL. A final regression analysis was performed to examine which aspects of sexual health were associated with QoL, adjusted for variables that were significant in the primary analysis among patients. Statistical analyses were performed in STATA v17.0 and significance was set at p≤0.05.

## Results

### Socio-demographic and clinical characteristics

From the 566 couples included in the eQuiPe study, 352 couples completed at least one questionnaire in the last 18 months of the patient’s life and were included in this analysis (Figure 1). Patients had a mean age of 66 years (SD 10) and 57% were male, and partners had a mean age of 65 years (SD 10) (Table 1). The most common primary tumors were lung (26%), colorectal (20%) and breast (12%). There were 4 same-sex couples, and most couples (98%) had a relationship for more than 5 years.

**Table 1.** Socio-demographic and clinical characteristics of the study population.

|  |  | Patients<br>N=352 |  | Partners<br>N=352 |  |
| --- | --- | --- | --- | --- | --- |
| Age (years) |  |  |  |  |  |
| Mean (SD), range |  | 65.6 (9.5) | 29-88 | 64.9 (10.0) | 19-87 |
|  |  | **n** | **%** | **n** | **%** |
| Sex | Male | 200 | 57% | 156 | 44% |
|  | Female | 152 | 43% | 196 | 56% |
| Religion | Yes | 232 | 66% | 223 | 63% |
|  | No | 118 | 34% | 128 | 37% |
| Educational level** | Low | 109 | 31% | 98 | 28% |
|  | Medium | 137 | 39% | 159 | 45% |
|  | High | 102 | 29% | 93 | 26% |
| Relationship duration | ≤5 years | 6 | 2% | 6 | 2% |
|  | >5 years | 346 | 98% | 346 | 98% |
| Children* | Yes | 282 | 80% | 300 | 85% |
|  | No | 44 | 13% | 50 | 14% |
|  | Missing | 26 | 7% | 2 | 1% |
| Children living at home*, ** | Yes | 46 | 13% | 50 | 14% |
|  | No | 236 | 67% | 248 | 70% |
|  | Missing | 70 | 20% | 54 | 15% |
| Primary tumor** | Lung | 92 | 26% | - | - |
|  | Colorectal | 72 | 20% | - | - |
|  | Breast | 41 | 12% | - | - |
|  | Prostate | 39 | 11% | - | - |
|  | Other | 107 | 30% | - | - |
| Received treatment in the last 18 months** | Yes | 311 | 88% | - | - |
|  | No | 40 | 11% | - | - |
| Comorbidities* | Yes | 220 | 63% | - | - |
|  | No | 109 | 33% | - | - |
|  | Missing | 23 | 7% | - | - |

\*Missing data were >5%.
\*\*Percentages do not always add up to 100 percent because of missings.

### Sexual health in patients and partners in the end of life

Patients sexual activity (range 15-19) and satisfaction (range 40-45) were relatively stable in the last 18 months of life (Figure 2). Sexual desire decreased in the last 18 months, from 18 in 16-18 months before death to 13 in the last 3 months respectively. Enjoyment was 31 (SD 33) 16-18 months before death and was 36 (SD 34) in the last 3 months of life. Mixed effect regression analysis confirmed that time to death was not associated with satisfaction, enjoyment or activity, only with a decrease in sexual desire (β 0.37, 95%CI 0.07-0.67) (Table 2).

**Figure 2.**
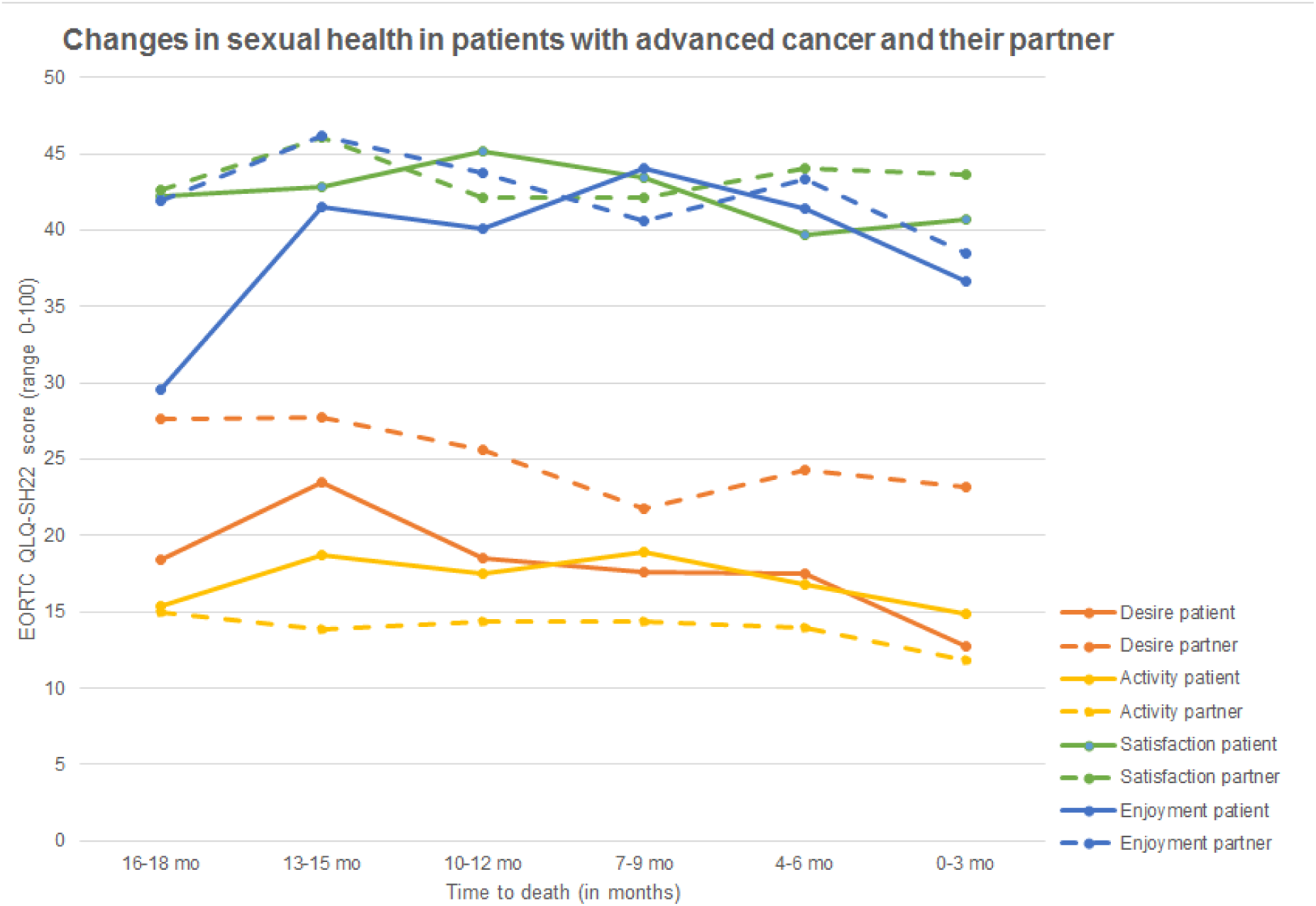
Sexual desire, activity, satisfaction, and enjoyment of patients with advanced cancer and their partner, in the last 18 months of life (n couples=352)

**Table 2.** Mixed-effects multivariable regression on the association between patient characteristics and sexual desire, activity, satisfaction, and enjoyment of patients.

|  | Desire<br>N=303 |  |  | Activity<br>N=303 |  |  | Satisfaction<br>N=303 |  |  | Enjoyment<br>N=169 |  |  |
| --- | --- | --- | --- | --- | --- | --- | --- | --- | --- | --- | --- | --- |
| | B | 95% CI | p-value | $\beta$ | 95% CI | p-value | $\beta$ | 95% CI | p-value | $\beta$ | 95% CI | p-value |
| Time to death | 0.37 | [0.07, 0.67] | **.017** | -0.07 | [-0.40, 0.26] | .695 | 0.08 | [-0.42, 0.57] | .753 | -0.44 | [-1.15, 0.26] | .219 |
| Sex, men | Ref |  |  |  |  |  |  |  |  |  |  |  |
| Women | -14.26 | [-19.73, -8.79] | **<.001** | -7.86 | [-13.22, -2.50] | **.004** | -0.13 | [-7.98, 7.73] | .974 | -10.70 | [-21.95, 0.55] | .062 |
| Age | -0.16 | [-0.42, 0.11] | .246 | -0.21 | [-0.46, 0.06] | .122 | -0.13 | [-0.51, 0.25] | .511 | -0.34 | [-0.88, 0.21] | .227 |
| Educational level, low | Ref |  |  |  |  |  |  |  |  |  |  |  |
| Medium | 0.10 | [-5.45, 5.6] | .973 | -0.36 | [-5.76, 5.05] | .897 | -0.02 | [-7.93, 7.90] | .996 | 10.34 | [1.19, 21.87] | .079 |
| High | -0.82 | [-7.02, 5.37] | .794 | -1.56 | [-7.59, 4.47] | .612 | 1.62 | [-7.20, 10.44] | .719 | 10.31 | [-2.62, 23.25] | .118 |
| Treatment, no | Ref |  |  |  |  |  |  |  |  |  |  |  |
| Treatment, yes | 1.35 | [-2.55, 5.25] | .497 | -1.40 | [-5.71, 2.91] | .524 | -1.67 | [-8.13, 4.80] | .613 | 0.78 | [-8.76, 10.32] | .873 |
| Comorbidities, no | Ref |  |  |  |  |  |  |  |  |  |  |  |
| Comorbidities, yes | -3.42 | [-8.37, 1.53] | .175 | -1.84 | [-6.66, 2.98] | .455 | 3.68 | [-.38, 10.75] | .307 | 2.68 | [-7.31, 12.68] | .599 |
| Physical functioning | 0.20 | [0.12, 0.28] | **<.001** | 0.19 | [0.10, 0.27] | **<.001** | 0.12 | [-0.01, 0.25] | .071 | 0.34 | [0.15, 0.53] | **.001** |
| Primary tumor, lung | Ref |  |  |  |  |  |  |  |  |  |  |  |
| Colorectal | -2.61 | [-9.62, 4.41] | .466 | -0.59 | [-7.43, 6.25] | .865 | 7.63 | [-2.41, 17.67] | .136 | -5.82 | [-19.55, 7.92] | .406 |
| Breast | 1.00 | [-7.57, 9.55] | .820 | 6.50 | [-1.83, 14.84] | .126 | 18.42 | [6.21, 30.62] | **.003** | 5.86 | [-11.17, 22.89] | .500 |
| Prostate | -20.26 | [-28.59, -11.92] | **<.001** | -10.85 | [-18.96, -2.74] | **.009** | -2.10 | [-13.99, 9.79] | .730 | -30.87 | [-50.91, 10.83] | **.003** |
| Other | -4.24 | [-10.25, 1.76] | .166 | 1.60 | [-4.27, 7.47] | .594 | 5.27 | [-3.33, 13.87] | .230 | -1.05 | [-13.21, 11.12] | .866 |
| Body image | 0.00 | [-0.36, 0.37] | .981 | -0.02 | [-0.41, 0.36] | .903 | -1.02 | [-1.58, -0.45] | **<.001** | -0.70 | [-1.52, 0.12] | .093 |
| QoL partner | 0.08 | [-0.03, 0.19] | .154 | 0.05 | [-0.06, 0.16] | .390 | 0.06 | [-0.11, 0.23] | .491 | 0.09 | [-0.14, 0.32] | .432 |

Partners sexual desire (range 22-28), activity (range 12-15), satisfaction (range 42-46), and enjoyment (range 39-46) remained relatively stable in the last 18 months of the patients’ life. Mixed effect regression analysis confirmed that time to death was not associated with sexual desire, activity, satisfaction, or enjoyment (Table 3).

**Table 3.**
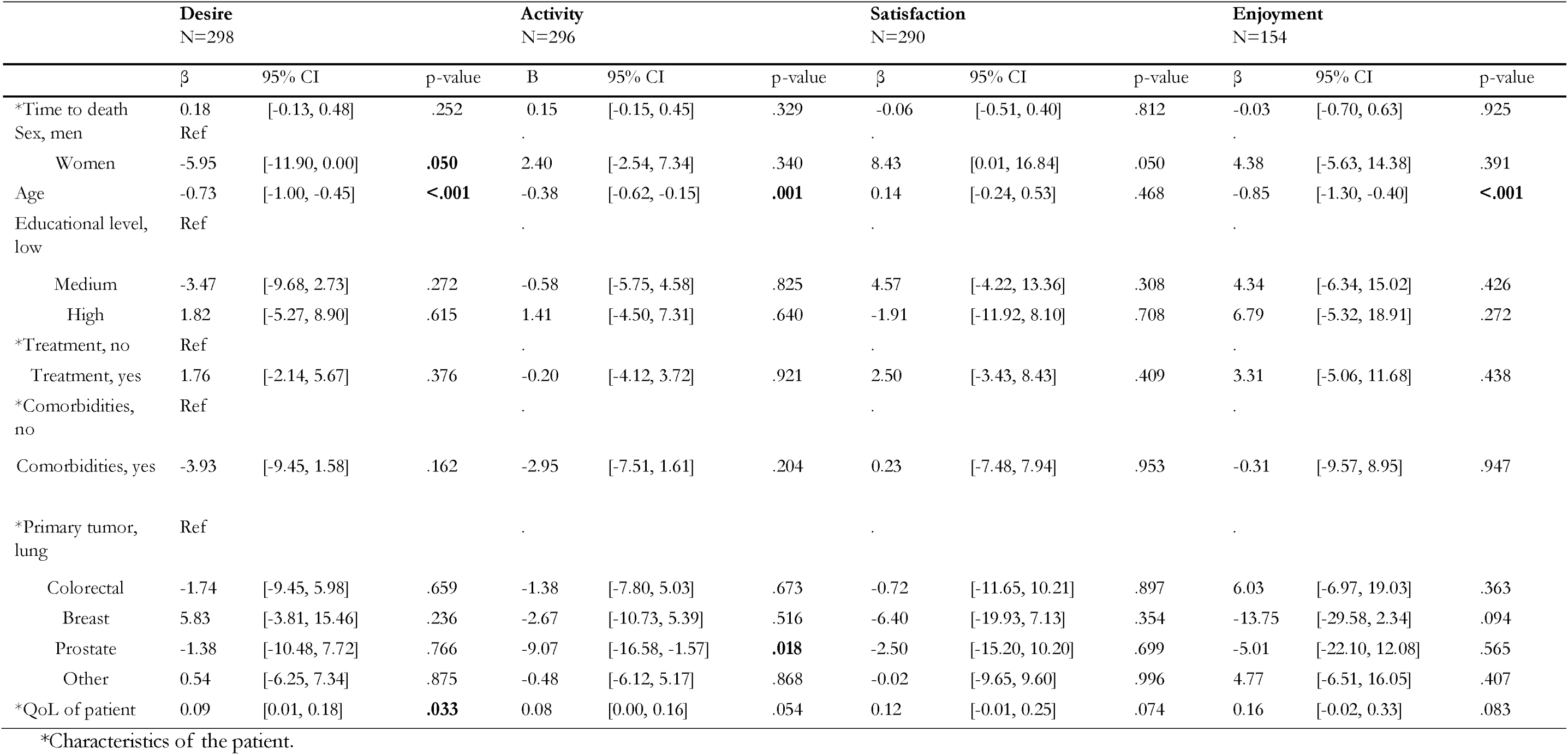
Mixed-effects multivariable regression on the association between partner and patient characteristics and sexual desir e, activity, satisfaction, and enjoyment of partners.

### Factors associated with sexual health in patients and partners at the end of life

Multivariable analyses showed that female patients had a steeper decline over time in sexual desire (β −14.26, p<0.05) and sexual activity (β –7.86, p<0.05) compared to male patients (Table 2). Better physical functioning was associated with higher scores on sexual desire in time (β 0.20, p<0.05), with higher levels of sexual activity (β 0.19, p<0.05) and sexual enjoyment for patients (β 0.34, p<0.05) (Table 2). Patients with a more positive body image also showed a significantly higher sexual satisfaction (β −1.02, p<0.05) compared to those with a lower body image.

In partners, multivariable analysis showed that female partners also reported significantly lower levels of sexual desire over time (β −5.95, p≤0.05), compared to male partners (Table 3). In partners, sexual desire (β −0.73, p<0.05), sexual activity (β −0.38, p<0.05) and sexual enjoyment (β −0.85, p<0.05) decreased with older age. Moreover, a better global QoL of the patient was associated with higher sexual desire in partners (β 0.09, p<0.05).

### Association between sexual health and QoL of patients over time

Sexual desire (β 0.11, p<0.05), sexual activity (β 0.08, p<0.05) and sexual satisfaction (β 0.06, p<0.05) were significantly associated with higher global QoL over time in patients (Table 4).

**Table 4.** Mixed-effects multivariable regression on the association between global QoL and sexual desire, activity, satisfaction, and enjoyment of patients *.

| Global QoL | N | $\beta$ | 95% CI | p-value |
| --- | --- | --- | --- | --- |
| Sexual desire | 316 | 0.11 | [0.04, 0.17] | **.001** |
| Sexual activity | 316 | 0.08 | [0.02, 0.14] | **.006** |
| Sexual satisfaction | 316 | 0.06 | [0.02, 0.10] | **.007** |
| Sexual enjoyment | 176 | 0.01 | [-0.06, 0.07] | .845 |

\*This model is corrected for time to death, age, sex, physical functioning, body image and primary tumor of the patient.

## Discussion

### Main findings

Couples confronted with advanced cancer reported diminished scores for sexual health in the end of life, and most aspects of sexual health remained stable over time for both patients and their partners. Only sexual desire in patients significantly decreased during the end of life. Patients’ sexual health was associated with physical functioning, tumortype and their partners’ age while partners’ sexual health was primarily associated with their own age. Moreover, sexual desire, activity, and satisfaction were significantly associated with global quality of life, indicating that these elements contribute to overall well-being in the last phase of life of patients with advanced cancer.

The sexual health scores of patients with advanced cancer in this study are partially consistent with previous research on sexual health. Normative data from Dutch cancer survivors indicated that 61% of individuals were not sexually active or only minimally so (37). This aligns with our study’s results, where both patients and partners exhibited modest scores in sexual health. Furthermore, the mean sexual satisfaction of patients in our study (42) was similar to that of a cross-cultural field study, where patients with recurrent or progressed cancer reported a mean score of 39 (38). However, normative data from the Netherlands and Norway indicate higher levels of sexual satisfaction compared to the findings of our study (37,39). The lower scores observed in our study suggest that patients with advanced cancer may experience reduced sexual satisfaction as a consequence of their illness.

Sexual enjoyment has not been widely explored before and comparing our findings with existing literature is difficult. However, available results on sexual enjoyment in patients with curative cancer show either similar or higher scores compared to the levels of enjoyment in our study in patients with advanced cancer (mean score 39) (17,25,40). The lower levels of sexual enjoyment in patients with advanced cancer suggest that patients may experienced more problemens with sexual enjoyment at the end of life than they do earlier in their (curative) cancer trajectory.

Sexual health is often assumed to change at the end of life. However, in our study most aspects of sexual health remained stable during the end of life, except for patient’ sexual desire, this decreased in time towards death. Research on sexual health underpins this finding that for most patients the need for sexuality remains important, however there is a shift in sexuality towards less physicality, and sexuality is redefined toward a deeper emotional connection at the end of life (10,13,26,41). This shift in sexuality may (partly) explain why patients and partners in our study are relatively satisfied with their sex life, despite lower scores in sexual activity.

Our study also showed that some patients’ sexual health was more affected than others. Patients with prostate cancer showed a stronger decrease in sexual desire, activity, and enjoyment, than patients with lung cancer. Also, partners showed a decrease in sexual activity if patients were diagnosed with prostate cancer. Previous research concluded that patients with prostate cancer may experience sexual dysfunction due to androgen deprivation therapy and its impact on daily functioning, caused by the disease or treatment (41–43). This sexual dysfunction likely directly affects multiple aspects of sexual health, leading to lower sexual health scores, as seen in this study. Moreover, female patients had a greater decline in sexual desire and sexual activity, and female partners also showed lower scores in sexual desire, compared to men. This could be explained by previous studies that highlighted increased emotional distress in women, regardless of role (patient or partner) (44,45), which may affect sexual health. Also, older partners showed a stronger decrease in sexual desire, activity, and enjoyment. An integrative review suggested that older patients with cancer are more likely to report decreased sexual health, in part due to barriers in seeking help caused by feelings of embarrassment and taboos (46,47). A higher age and decreased sexual activity are consistent with data from the general population in the United States (48). However, age did not affect sexual health in patients in our study, only of partners.

Decreased physical functioning of patients was also associated with a greater decline in multiple aspects of sexual health, including sexual desire, activity, and enjoyment. This is in line with a previous study in patients with metastatic cancer that showed that more than half of the patients reported that poor physical condition negatively affected their sex life (6). This can be explained by the fact that when patients function worse physically, it negatively contributes to their QoL and therefore they experience more barriers to engaging in sexual interactions. Furthermore, most patients in this study received treatment, and it is known that the severity and intensity of treatment could affect the patient’s QoL (49), which can lead to decreased sexual health. We also showed that sexual desire, activity, and satisfaction were associated with a better overall QoL, indicating that sexual health remains an important element of QoL for patient at the end of life and their partners. This is in line with prior research, as patients reported that sexual health remained a priority, even at the end of life (8,10).

### Study limitations

Some limitations of the study need to be addressed. Patient recruitment was performed by the attending physician, a process that may have contributed to selection bias, this could lead to an overestimation of sexual health. The EORTC’s sexual health questions are not widely used yet, which hampers the comparison of the results of this study. Also limited information regarding normative values are present of the sexual health items. Lastly, this study focused exclusively on couples to fill a gap in literature where the primary emphasis has been on patients only. However, a recent article has underscored that individuals who do not have a partner also deal with obstacles regarding intimacy and sexuality when dealing with terminal illness (50).

### Clinical implications

This study underpins the importance to discuss the topic of sexual health in the end of life. Multiple studies showed that most patients with a life-threatening disease feel the need to discuss challenges in sexuality and intimacy (8,14). More than 75% of oncology healthcare providers agreed that it is their responsibility to initiate discussions about sexuality and intimacy (18,51,52). Nevertheless, several studies concluded that healthcare providers often fail to initiate these discussions (10,19,21,53,54). Barriers to address sexual health include feelings of discomfort, inadequate training, time constraints, assumptions about elderly patients and sexuality, and limited understanding of intimate relationship dynamics in the context of metastatic cancer (8,18,21,55). Patients are also reluctant to discuss these topics due to feelings of embarrassment or because they expected healthcare providers to take the lead (10,13). Further research is essential to examine whether patients perceive decreased sexual health as a concern both prior to diagnosis and at the end of life, as well as to assess their interest in discussing sexual health with their healthcare professional. The stepwise Permission Limited Information Specific Suggestions Intensive Therapy (PLISSIT) model is a tool that proved to be useful among healthcare providers and has a positive effect on QoL and sexual functioning in both patients and partners (56).

## Conclusion

Sexual health in patients with advanced cancer and their partners remains relatively stable in the last 18 months of life, only sexual desire in patients decreases in time towards death. Patients with prostate cancer, female patients, and older partners are more likely to experience lower sexual health. Patients with a decreased physical functioning are also more likely to have a stronger decline in most aspects of sexual health and lower sexual health is associated with poorer QoL. Maintaining or improving QoL is paramount for patients with incurable cancer and their partners. Therefore, addressing sexual health in couples confronted with incurable cancer is important, also within the context of palliative care.

## Data Availability

All data produced in the present study are available upon reasonable request to the authors

https://gegevensaanvraag.iknl.nl/gegevensaanvraag

## Aknowledgements

We extend our gratitude to all patients and partners who took part in the eQuiPe study, as well as to the NCR for their support.

## Declarations

### Authorship

**Isabel S. van der Meer**: formal analysis, methodology, project administration, resources, visualization, and writing – original draft. **Moyke A.J. Versluis**: formal analysis, methodology, project administration, and supervision. **Natasja J.H. Raijmakers**: funding acquisition, methodology, project administration, validation and supervision. All authors provided feedback in writing and conceptualization – editing and reviewing.

### Research ethics and patient consent

All patients and partners gave written consent to participate in the eQuiPe study.

### Data management and sharing

Data from the eQuiPe study, formatted in DDI 3.x XML, can be retrieved through Questacy and is hosted on the PROFILES registry (Patient-Reported Outcomes Following Initial Treatment and Long-term Evaluation of Survivorship) at www.profilesregistry.nl. To uphold rigorous standards for enduring data preservation and accessibility, the study adhered to the principles outlined in the “Data Seal of Approval” created by Data Archiving and Networked Services (DANS) (www.datasealofapproval.org).

**Appendix 1:** In- and exclusion criteria for patients and partners.

|  | *Patients* | *Partners* |
| --- | --- | --- |
| *Inclusion criteria* | Patients with prostate cancer were included when they had metastases, and their cancer was castrate-resistant. | - |
|  | Patients with breast cancer were included when their metastases were located in multiple organ systems. | - |
|  | Patients who were able to complete the questionnaire in Dutch | Partners who were able to complete the questionnaire in Dutch. |
| *Exclusion criteria* | Patients with breast or prostate cancer with a relative good diagnosis. | - |
|  | Patients with severe mental illness. | - |
|  | Patients younger than 18 years. | Partners younger than 18 years. |

